# A Transparent Four-Feature Logistic Model for Depression Screening in Assisted-Living Facilities

**DOI:** 10.1101/2025.07.14.25331539

**Authors:** Kevin Mekulu, Faisal Aqlan, Hui Yang

## Abstract

Depression in older adults is both common and frequently underdiagnosed, especially in assisted-living communities, where it often co-occurs with mild cognitive impairment (MCI), creating a complex and vulnerable clinical landscape. Despite this urgency, scalable, interpretable, and easy-to-administer tools for early screening remain scarce. In this study, we introduce a transparent and lightweight AI-driven screening model that uses only four linguistic features extracted from brief conversational speech, to detect depression with high sensitivity. Trained on the DAIC-WOZ dataset and optimized for deployment in resource constrained settings, our model achieved strong discriminative performance (AUC = 0.760) with a clinically calibrated sensitivity of 92%. Beyond raw accuracy, the model offers insights into how affective language, syntactic complexity, and latent semantic content relate to psychological states. Notably, one semantic feature derived from transformer embeddings, *emb_1*, appears to capture deeper emotional or cognitive tension not directly expressed through lexical negativity. We propose this component as a potential digital biomarker of cognitive-affective strain, warranting further longitudinal study. Our approach outperforms many more complex models in the literature, yet remains simple enough for real-time, on-device use, marking a step forward in making mental health AI both interpretable and clinically actionable.

## Introduction

It often begins in quiet ways: a softening of tone, a hesitation mid-sentence, a drift toward vagueness or avoidance. Depression in older adults is among the most underrecognized challenges in clinical care, particularly within assisted-living communities, where mental health resources are often limited, and symptoms may be mistaken for normal aging. What makes the situation more urgent is that depression rarely occurs in isolation. Mild cognitive impairment (MCI) frequently co-occurs with depressive symptoms in this population, creating a dual burden that increases the risk of progression to dementia, functional decline, and hospitalization^1,2^.

Despite this high-stakes context, routine depression screening in eldercare remains inconsistently implemented. Traditional tools like the PHQ-9 or Geriatric Depression Scale require dedicated time, trained personnel, and patient literacy—and even then, they may miss early or masked presentations of mood disturbance. Furthermore, most screening instruments offer limited visibility into the cognitive underpinnings of emotional change, overlooking subtle linguistic cues that may reflect both affect and executive strain.

Recent advances in artificial intelligence (AI) and natural language processing (NLP) offer a promising way forward. By analyzing not only what individuals say, but how they say it, we can extract early markers of both mood and cognition. Yet many state-of-the-art AI models—particularly deep learning architectures—come with a trade-off: high performance at the expense of interpretability. In clinical settings, where stakes are high and trust is critical, such opacity can hinder adoption and raise ethical concerns^3,4^.

Our goal in this study was to build a screening tool that doesn’t force this trade-off. We present a fully interpretable, four-feature model that screens for depression using only a short speech sample. Unlike acoustic systems—which require clean audio and specialized hardware—our model operates on text transcripts, making it easier to deploy in real-world care settings where background noise, privacy, and technical variability present real barriers^5,6^.

Trained on the DAIC-WOZ dataset^7–9^, our model uses three handcrafted linguistic features and one derived semantic embedding. Each is interpretable, lightweight, and grounded in either psycholinguistic theory or observed speech dynamics. Most notably, a single reduced embedding dimension—*emb_1*—emerged as a strong predictor. Through qualitative analysis, we found that this dimension may capture latent psychological conflict or cognitive-affective strain, even in the absence of overtly negative language. This raises the possibility of a new class of semantic biomarkers for affective and cognitive health—a hypothesis we believe warrants further study.

In the sections that follow, we describe our modeling approach, evaluate performance against peer systems, and explore how this transparent, text-only pipeline could support timely, scalable, and clinically meaningful screening for depression in aging populations.

## Results

From the beginning, our aim was clear: to build a model that not only performs well but also tells a story—about the speaker, about their state of mind, and about the linguistic markers that quietly signal emotional distress. Rather than relying on dozens of features or black-box architectures, we sought simplicity without compromise.

After preprocessing and extracting participant-only speech, our dataset consisted of 188 valid samples: 106 for training, 35 for development, and 47 for held-out testing. Using sparse logistic regression with an *𝓁*_1_ penalty, the model distilled this data into just four features: a depression-focused word ratio (neg_words_r), a VADER-based sentiment score (sent_neg), a syntactic complexity metric (avg_dep_dist), and a single, reduced semantic component (emb_1) from a sentence-transformer embedding. These four alone were sufficient to tell the story.

The final model, trained with *C* = 0.6310 and calibrated for high sensitivity, achieved an AUC of 0.760 on the test set. At the fixed threshold of 0.170, selected to ensure sensitivity*≥* 0.92 on the development set, the model reached 92% sensitivity and 57% specificity—prioritizing the detection of true cases, as warranted by our clinical setting. Figure 1 presents the ROC curve on the test set, reflecting consistent discriminative performance.

**Figure 1.**
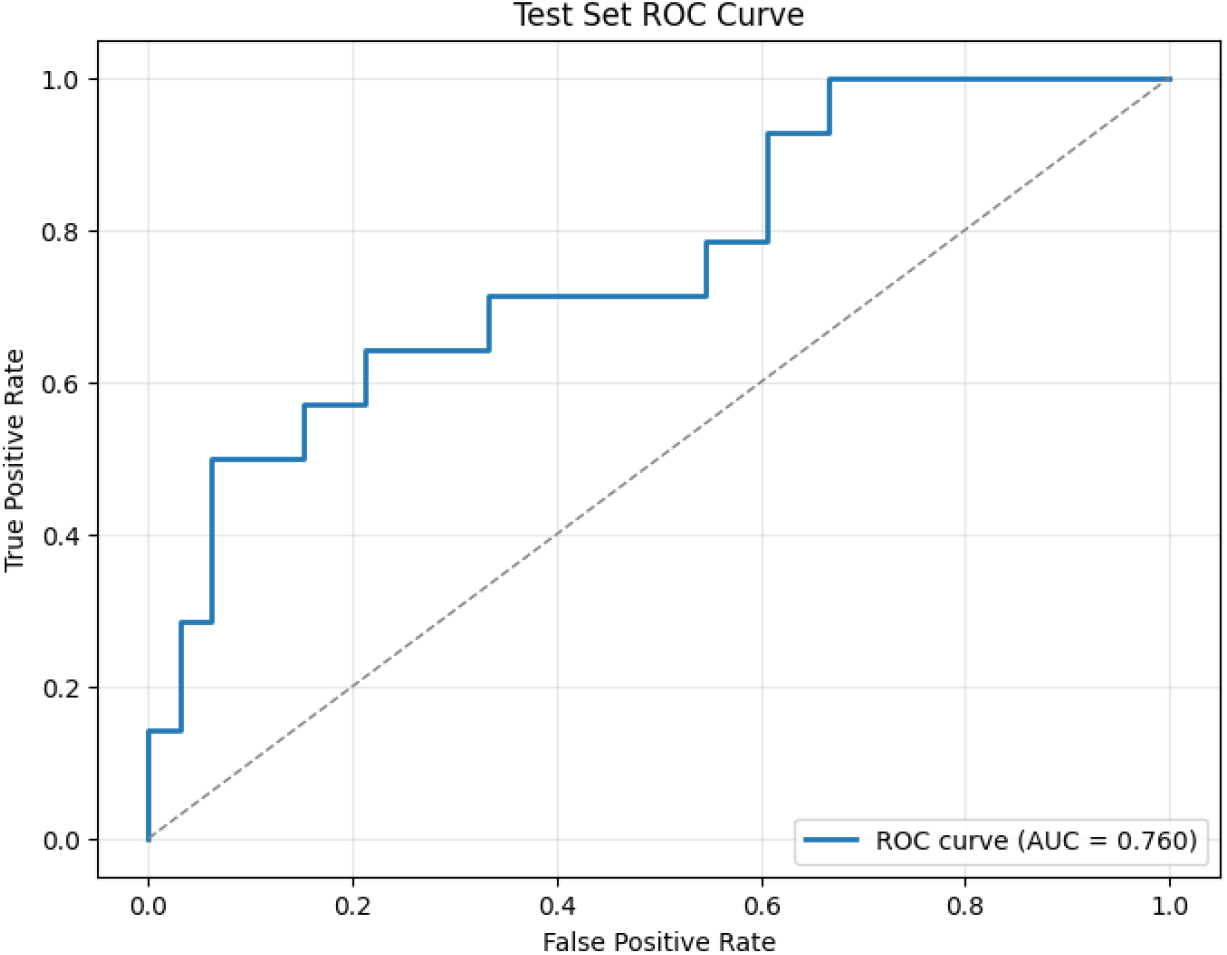
Test set ROC curve for our four-feature depression screening model. The model achieves an AUC of 0.760 on held-out data, with a clinically calibrated threshold (marked) yielding a sensitivity of 92%. This operating point reflects our design priority: maximizing true positive detection in high-risk settings like assisted living, where missed cases of depression can have serious downstream consequences. Despite its simplicity, the model maintains strong discriminative power—comparable to more complex systems—while offering full interpretability and real-world deployability.

### Interpreting the Model: What the Features Tell Us

Each of the four retained features carries distinct weight in shaping the model’s predictions—and each reflects a different layer of how depression can manifest in spontaneous language.

| Feature | Coefficient | Absolute Value |
| --- | --- | --- |
| neg_words_r | 0.458 | 0.458 |
| emb_1 | 0.295 | 0.295 |
| sent_neg | 0.275 | 0.275 |
| avg_dep_dist | -0.124 | 0.124 |

The two affective features—`neg_words_r` and `sent_neg`—provide a lexical and sentiment-based signal, respectively, both of which are well-established in prior work on depression detection. Participants who used more negative-valence language, or who expressed sadness or hopelessness explicitly, were more likely to be flagged by the model.

But the third feature, `avg_dep_dist`, opens a different window. This metric captures the syntactic complexity of participants’ speech: how far apart words are, on average, from the other words that govern them grammatically. Greater distances tend to reflect more nested, elaborate sentence structures—while shorter distances suggest simpler, flatter syntax. In the context of geriatric mental health, syntactic simplification has been observed in both depressive disorders and early cognitive decline^10,11^. Thus, we interpret this feature not just as a linguistic proxy, but as a possible marker of reduced working memory or executive function. While speculative, it raises the possibility that even in speech ostensibly about mood, subtle signs of cognitive strain are already embedded.

### Unpacking emb_1: Latent Meaning in Speech

The most intriguing signal, however, came from `emb_1`—a single semantic dimension extracted via SVD from a 384-dimensional transformer-based embedding. Unlike the handcrafted features, this one was data-driven, abstract, and not tied to any single word or phrase. Yet it proved nearly as influential as the lexicon-based features.

To understand why, we manually examined transcripts with the highest and lowest values of `emb_1`. High-scoring interviews often included affectively loaded reflections, references to uncertainty, or socially fraught topics—such as systemic injustice or personal distress. For example:

**TRANSCRIPT 426 (high emb_1)**: *“*…*the weather, the women, the opportunities—that’s about it. Racism, police brutality, injustice, that’s it*… *“*

**TRANSCRIPT 320 (high emb_1)**: *“*…*yes, I’m a little nervous*… *what do I do now? I don’t know right now*… *“* By contrast, low `emb_1` scores were associated with routine, fact-based, emotionally neutral language:

**TRANSCRIPT 480 (low emb_1)**: *“*…*New York City*… *once a year*… *both urban*… *east coast has a little bit maybe a little more culture*… *“*

**TRANSCRIPT 450 (low emb_1)**: *“*…*born and raised*… *I love the ocean*… *I love the hiking*… *“*

What emb_1 appears to capture, then, is a kind of semantic gravity—a dimension of language that reflects latent psychological weight even when overtly “depressive” words are absent. This observation suggests a testable hypothesis: that semantically derived features like `emb_1` may serve as digital biomarkers of cognitive-affective conflict, where speech reveals internal tension not yet crystallized into clinical symptoms. Future work could explore whether shifts in this component predict transitions into depressive episodes or the early stages of MCI.

## Discussion

In assisted-living communities, where cognitive and emotional vulnerabilities often converge, the early detection of depression is more than a screening challenge—it’s a clinical imperative. Depression not only worsens functional outcomes in aging populations, but also frequently coexists with conditions like mild cognitive impairment (MCI), obscuring the clinical picture and delaying intervention^1,2^. Our goal was to create a screening tool tailored for this environment—one that is interpretable, low-burden, and built to earn trust at the point of care.

What we found was both validating and unexpected. Our sparse, four-feature model delivered strong performance (AUC = 0.760) using only a brief segment of participant speech, without relying on acoustic signals or deep black-box architectures.

But beyond its numbers, the model offers insights into how depression subtly shapes language—and how those patterns can be captured and interpreted in ways that are clinically meaningful.

One such insight emerged from the inclusion of syntactic complexity, measured via average dependency distance. While simple in design, this feature draws from well-established psycholinguistic theory: reductions in grammatical complexity and clause nesting have been linked to both depression and early cognitive decline^10,11^. In our model, flatter syntactic structures were associated with higher depression risk—a finding that, while not diagnostic on its own, may reflect diminished working memory or reduced executive function during speech planning.

Perhaps the most surprising result came from `emb_1`, a semantic dimension derived from a sentence-transformer embedding. Despite being abstract and data-driven, this component consistently aligned with expressions of unease, self-doubt, or affective weight—even when overtly “negative” language was absent. This opens a provocative hypothesis: that semantically derived features like `emb_1` may serve as early indicators of latent cognitive-affective strain. If validated in future longitudinal work, this could mark the beginning of a new class of digital biomarkers—one capable of detecting risk before traditional screeners do. Importantly, the strength of this approach lies not only in its interpretability, but also in its readiness for real-world use. As shown in Figure 2, the full deployment pipeline—from speech capture to risk flag—is lightweight, modular, and compatible with existing care workflows. This positions the tool not just as a research model, but as a scalable solution for routine screening in assisted-living communities.

**Figure 2.**
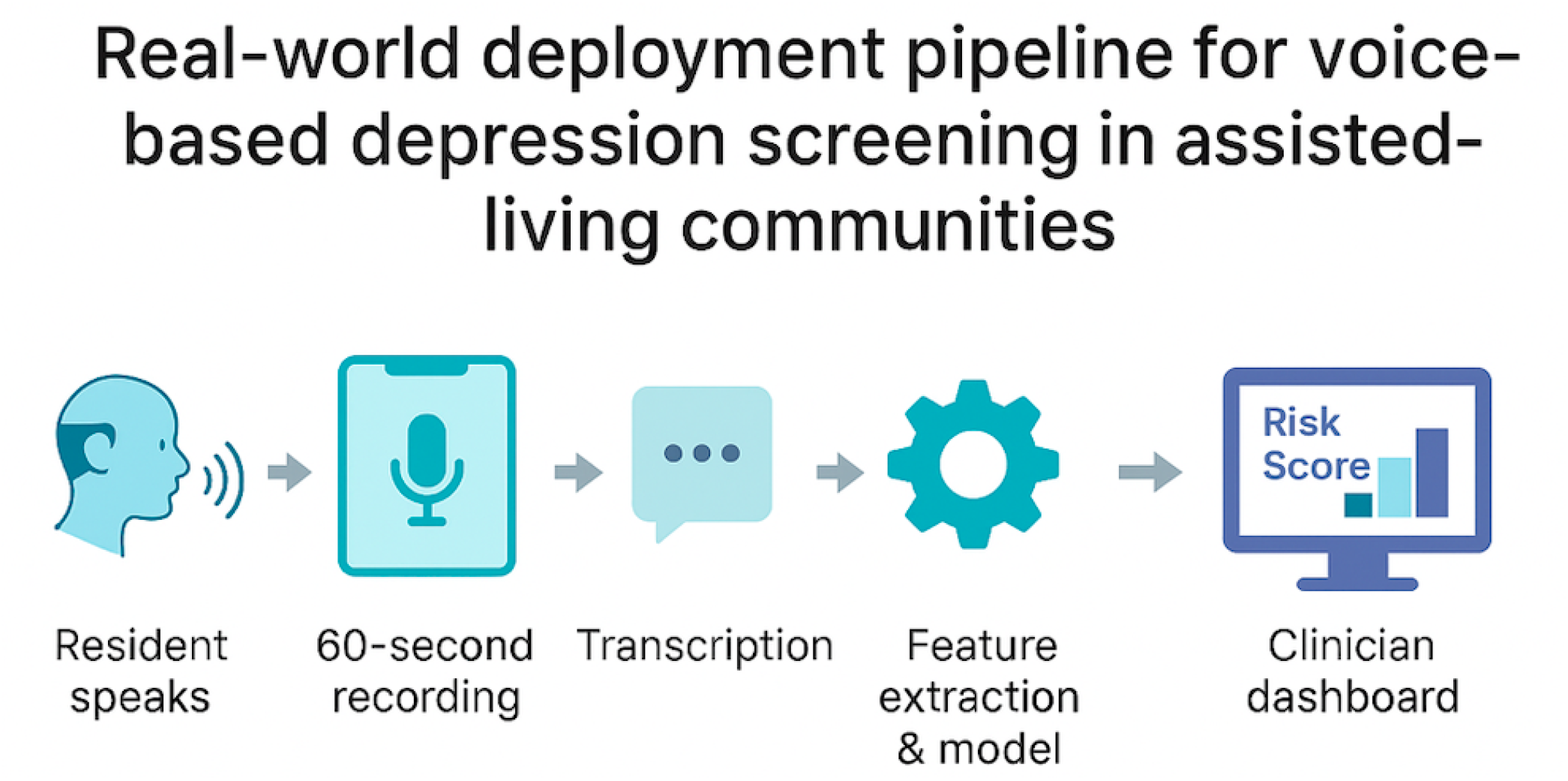
Real-world deployment pipeline for voice-based depression screening in assisted-living communities. A 60 second speech sample is recorded on a standard device, transcribed, and processed through a four-feature logistic regression model. The resulting risk score and feature explanations are displayed on a clinician dashboard for immediate review and follow-up.

### Limitations and Future Directions

While our results are promising, several limitations must be acknowledged. First, the model was trained and evaluated on the DAIC-WOZ corpus—a structured research dataset with a standardized interview protocol. Although widely used in depression detection research, this setting may not fully reflect the linguistic variability, cultural diversity, or conversational context found in real-world assisted-living environments. Future validation on naturalistic data from clinical or residential care settings is essential.

Second, our model currently relies solely on text-based features, omitting acoustic, visual, and interactional cues that could further enrich screening accuracy. While this choice was intentional—favoring deployability and robustness to noise—it also means we may be missing signals captured by multimodal systems.

Third, the semantic embedding feature (`emb_1`) offers strong predictive value but remains abstract. Although our qualitative analysis suggests it captures cognitive-affective tension, its exact linguistic and neuropsychological underpinnings remain speculative. Future work should explore whether such embeddings correlate with clinical assessments, neuroimaging findings, or longitudinal outcomes—particularly in individuals at risk for both depression and cognitive decline.

Finally, while our model was optimized for high sensitivity, this choice comes with a trade-off: reduced specificity. In practice, this may lead to some false positives. However, given the relatively low cost of follow-up evaluation in most assisted-living contexts—and the high cost of missed cases—we believe this trade-off is clinically justified.

Moving forward, we aim to test this model in real-world deployments, integrate it with structured cognitive tasks such as verbal fluency, and explore its utility in longitudinal screening for MCI and dementia. We also envision adapting the system for multilingual use and evaluating its fairness across gender, race, and educational background.

## Methods

### Overview

Our aim from the outset was to design a screening pipeline that could operate not just in theory, but in the messy, time-constrained, and resource-limited context of real-world elder care. That meant simplifying everything—from data inputs to model architecture—while still preserving the scientific rigor needed to make this a clinically trustworthy tool. Figure 3 summarizes our streamlined, four-stage workflow: from spoken interview to depression risk flag, all within a transparent and interpretable framework.

**Figure 3.**
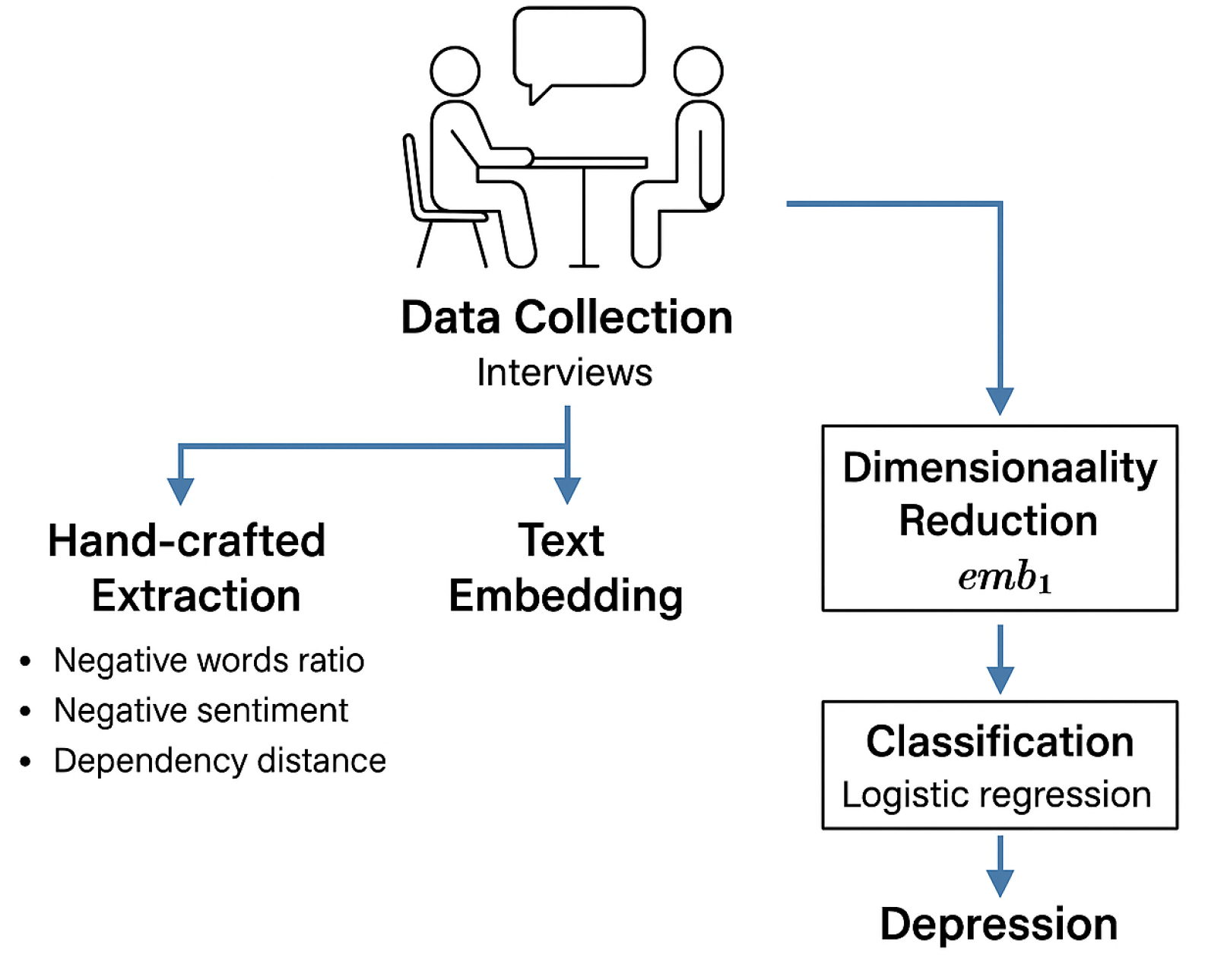
Workflow diagram for our interpretable depression screening model. Participant interviews serve as the input, followed by parallel extraction of handcrafted linguistic features and text embeddings. Embedding vectors are reduced to a single dimension, and all features are passed to a sparse logistic regression classifier.

### Data Source and Preprocessing

We used the Distress Analysis Interview Corpus – Wizard of Oz (DAIC-WOZ) dataset^7^,^8^, a benchmark corpus released as part of the AVEC 2017 Challenge^9^. The dataset includes transcribed semi-structured clinical interviews between participants and a virtual interviewer, with depression severity labeled based on PHQ-8 scores^12^. Each participant completed the interview in a consistent format, and binary depression labels were derived using a cutoff on PHQ-8 total scores.

To ensure that our model would generalize to real-world clinical settings—and not simply learn the dialogue structure or cues from the interviewer—we filtered out all non-participant speech. The remaining transcripts were normalized: lowercased, stripped of punctuation, and cleaned of formatting inconsistencies. After filtering, we retained 188 usable samples: 106 for training, 35 for development, and 47 for held out testing.

### Feature Extraction

We focused on capturing three types of signals: affective language, sentence complexity, and latent semantic content.

#### Handcrafted features

Inspired by clinical language markers, we designed three interpretable metrics:

- **Negative word ratio (neg_words_r)** — The proportion of words in a transcript that match a curated depression lexicon (e.g., “hopeless,” “afraid,” “useless”).
- **Negative sentiment (sent_neg)** — A scalar value computed using the VADER sentiment analysis tool, capturing the overall negative valence of the text.
- **Average dependency distance (avg_dep_dist)** — A structural feature based on syntactic parsing, reflecting the average distance between tokens and their grammatical heads.

#### Text embeddings

Beyond surface language and structure, we aimed to capture semantic nuance. We encoded each transcript using the “all-MiniLM-L6-v2” sentence-transformer, a lightweight transformer optimized for performance on small devices. We then reduced the high-dimensional embeddings using truncated Singular Value Decomposition (SVD), and selected the second principal component—*emb_1*—based on exploratory correlations with depression status.

### Model Training and Calibration

We trained a logistic regression model with *𝓁*_1_ regularization (lasso) to enforce sparsity and interpretability. This approach allowed the model to ignore any features that didn’t meaningfully contribute to its predictions, yielding a final configuration with exactly four nonzero weights. We chose the optimal regularization strength (*C* = 0.6310) through grid search on the training set.

Probabilities from the model were calibrated using a sigmoid transformation. Then, on the development set, we selected a decision threshold that guaranteed high sensitivity—specifically, at least 92%. This resulted in a final threshold of 0.170, which was locked in before evaluating on the test set.

### Evaluation Strategy

Consistent with our clinical priorities, we focused on three primary metrics for model evaluation:

- **Area Under the Curve (AUC)** — to assess the model’s ability to discriminate between depressed and non-depressed participants across thresholds.
- **Sensitivity (Recall)** — to measure the proportion of truly depressed participants that were correctly flagged by the model.
- **Specificity** — to understand the model’s ability to avoid false positives.

These metrics were computed exclusively on the held-out test set (n = 47) using the fixed high-sensitivity threshold selected during development.

## Conclusions

This study demonstrates that depression in older adults can be effectively screened using only a short transcript and a transparent, four-feature model. By grounding each feature in linguistic theory and clinical relevance, we offer not just a tool, but a framework—one that balances interpretability with performance and is built for real-world use. Most notably, our analysis of a single semantic embedding dimension, `emb_1`, suggests the potential for new digital biomarkers that reflect latent emotional or cognitive tension even when traditional signals are absent. As aging populations grow and mental health needs in assisted-living communities intensify, tools like this—low-burden, explainable, and clinically aligned—will be essential. Future work will focus on validating this model in naturalistic settings, integrating it with structured cognitive tasks, and testing whether its semantic markers anticipate broader cognitive changes. The opportunity is clear: by listening closely to how people speak, we may detect distress long before it’s visible on a checklist.

## Data Availability

The dataset used in this study is the Distress Analysis Interview Corpus - Wizard of Oz (DAIC-WOZ), part of the AVEC 2017 Challenge. Access to the dataset is publicly available upon request for research purposes. Researchers can obtain the data by registering and agreeing to the data usage terms at the official repository: https://dcapswoz.ict.usc.edu/

## Acknowledgements

The authors of this work would like to acknowledge the NSF grants IIS-2302834 and MCB-1856132 for funding this research. Any conclusions found in this paper originate from the authors and do not necessarily reflect the views of the sponsor.

## Author contributions statement

K.M. conceived the research, performed analysis, and wrote the manuscript. F.A. & H.Y. provided feedback and reviewed the manuscript.

## Additional information

## References

1. Petersen, R. C. Mild cognitive impairment. The New Engl. J. Medicine 364, 2227–2234 (2011).

2. Mitchell, A. J., Vaze, A. & Rao, S. Clinical diagnosis of depression in primary care: a meta-analysis. The Lancet 374, 609–619 (2009).

3. Lipton, Z. C. The mythos of model interpretability. Queue 16, 31–57 (2018).

4. Caruana, R. et al. Intelligible models for healthcare: Predicting pneumonia risk and hospital 30-day readmission. In Proceedings of the 21th ACM SIGKDD International Conference on Knowledge Discovery and Data Mining, 1721–1730 (2015).

5. Balagopalan, A. & et al. Using natural language processing to screen for depression in primary care. NPJ Digit. Medicine 4, 1–9 (2021).

6. Alhanai, T., Ghassemi, M. & Glass, J. Detecting depression with audio/text sequence modeling of interviews. In Proc. Interspeech, 1716–1720 (2018).

7. Gratch, J. et al. The distress analysis interview corpus of human and computer interviews. In Proceedings of the Ninth International Conference on Language Resources and Evaluation (LREC’14), 3123–3128 (2014).

8. DeVault, D. et al. SimSensei Kiosk: A virtual human interviewer for healthcare decision support. In Proceedings of the 13th International Conference on Autonomous Agents and Multiagent Systems (AAMAS’14) (Paris, France, 2014).

9. Ringeval, F. et al. Avec 2017: Real-life depression, and affect recognition workshop and challenge. In Proceedings of the 7th Annual Workshop on Audio/Visual Emotion Challenge, 3–9 (ACM, 2017).

10. Roark, B., Mitchell, M., Hollingshead, K. et al. Spoken language derived measures for detecting mild cognitive impairment. IEEE Transactions on Audio, Speech, Lang. Process. 19, 2081–2090 (2011).

11. Frazier, M. et al. Speech-based markers for cognitive and psychiatric diagnosis. Curr. Opin. Psychol. 45, 101305 (2022).

12. Kroenke, K. et al. The phq-8 as a measure of current depression in the general population. J. Affect. Disord. 114, 163–173 (2009).

